# Clinical characteristics and associated factors of de novo and recurrent prostate cancer after kidney transplantation

**DOI:** 10.64898/2026.07.30.26359389

**Authors:** Kunle Apanisile, Meng-Hao Li, Giovanni Faddoul, Obi Ekwenna, Naoru Koizumi

## Abstract

Kidney transplant recipients experience a higher burden of several malignancies, yet the factors associated with prostate cancer presentation after transplantation remain poorly understood. Unlike malignancies strongly associated with impaired immune surveillance, prostate cancer has not consistently demonstrated an increased incidence after transplantation, suggesting that different mechanisms may underlie disease presentation. This study evaluated recipient, donor, transplant, immunologic, and immunosuppressive factors associated with prostate cancer phenotype after kidney transplantation.

A retrospective cohort study was conducted using national transplant registry data from adult kidney transplant recipients diagnosed with post-transplant prostate cancer between 2015 and 2024. Cases were classified as de novo (no pre-transplant history of prostate cancer) or recurrent (documented pre-transplant history). Multivariable Firth penalized logistic regression was used to evaluate factors associated with recurrent phenotype. Prespecified sensitivity analyses included deceased donor restricted models, incorporation of donor organ quality variables, and adjustment for time from transplantation to cancer diagnosis. Exploratory machine learning analyses included elastic net logistic regression, random forest, and extreme gradient boosting.

The cohort included 660 recipients, of whom 623 (94.4%) had de novo disease and 37 (5.6%) had recurrent disease. Recipient age was the only variable consistently associated with recurrent phenotype across primary and sensitivity analyses (adjusted odds ratio per year 1.11, 95% CI 1.05–1.17; p<0.001). Immunosuppressive regimen, donor characteristics, immunologic variables, and time from transplantation to cancer diagnosis were not independently associated with phenotype in the primary cohort. In deceased donor restricted analyses, alemtuzumab induction showed an exploratory association with recurrent phenotype, although estimates were imprecise. Machine learning models demonstrated modest discrimination and calibration and did not outperform penalized regression approaches.

These findings suggest that, among kidney transplant recipients with prostate cancer, differences between recurrent and de novo presentation are more closely associated with recipient age and underlying disease characteristics than with transplant exposures or specific immunosuppressive regimens.

## Introduction

Kidney transplantation remains the treatment of choice for patients with end-stage kidney disease, offering substantial survival and quality of life benefits compared with long-term dialysis [1]. These benefits, however, are accompanied by important long-term complications. Among these, post-transplant malignancy has emerged as a leading cause of late morbidity and mortality [2–4]. Kidney transplant recipients experience a markedly increased risk of cancer compared with the general population. This excess risk is largely attributable to chronic immunosuppression, impaired immune surveillance, oncogenic viral infections, and factors related to end-stage kidney disease and prolonged dialysis exposure [5–8]. Importantly, this excess cancer risk is not uniform across tumor types but reflects distinct biological pathways, with some malignancies strongly linked to immune dysregulation, whereas others, including prostate cancer, appear less consistently associated with immunosuppression.

The spectrum of malignancies observed after kidney transplantation differs from that of the general population. Cancers driven by infection and immune dysfunction, including post-transplant lymphoproliferative disorder, Kaposi sarcoma, and non-melanoma skin cancers, occur at disproportionately higher rates among transplant recipients [7–10]. In contrast, the epidemiology of prostate cancer in kidney transplant recipients is more heterogeneous. Prostate cancer is one of the most common malignancies among men globally, and its risk is strongly associated with advancing age and inherited susceptibility, although non-genetic factors, including obesity, dietary factors, and environmental exposures, may also contribute [11–16]. Meta-analyses of large transplant cohorts have not demonstrated a consistent increase in prostate cancer incidence after transplantation [17,18], a finding supported by national registry studies including a Swedish cohort [19–21]. At the same time, several cohort studies have reported higher crude incidence rates of prostate cancer among kidney transplant recipients, which are more plausibly explained by differences in age distribution, screening practices, and detection patterns rather than a true increase in biological risk [22–26]. Collectively, these observations suggest that analyses based solely on incidence may not fully capture the heterogeneity of prostate cancer presentation after transplantation.

A phenotype approach offers a more focused way to examine this heterogeneity. In clinical practice, de novo prostate cancer after transplantation and recurrence of pre-existing disease raise different questions. De novo disease may reflect recipient aging, screening intensity, and post-transplant exposures, whereas recurrent disease may reflect the biology of a prior malignancy, timing of transplantation, and the effect of immunosuppression on residual or previously treated disease. Distinguishing these presentations is therefore important, not because it estimates the absolute risk of prostate cancer or recurrence in the broader transplant population, but because it may identify whether affected recipients with recurrent disease differ meaningfully from those with de novo disease. Transplant oncology literature recognizes that pre-existing malignancy and de novo post-transplant cancer may carry different clinical implications and outcomes [27,28]. This distinction supports the evaluation of prostate cancer phenotypes as related but clinically separate entities rather than combining all post-transplant prostate cancer cases into a single outcome. Such characterization may help clarify whether recurrent prostate cancer in kidney transplant recipients is primarily associated with baseline patient factors, transplant factors, immunologic factors, or immunosuppression exposure.

To date, no large study using national transplant registry data has systematically compared the recipient, donor, transplant, immunologic, and immunosuppressive factors associated with de novo versus recurrent prostate cancer among kidney transplant recipients. In this context, the present study uses a national transplant registry cohort to compare these two phenotypes. The objective is to identify factors associated with prostate cancer phenotype using a multivariable analytical framework and to characterize how these factors differ within this population.

## Methods

### Study design and data source

This study was conducted as a retrospective cohort analysis using de-identified data from the United Network for Organ Sharing (UNOS) national transplant registry. The data were accessed for research purposes on 25 March 2026. The analytic cohort consisted of adult kidney transplant recipients who developed prostate cancer following transplantation between 2015 and 2024. The registry provided detailed information on recipient characteristics, donor factors, transplant procedures, immunosuppression, and clinical outcomes. The authors did not have access to information that could identify individual participants during or after the study. This study was reported in accordance with the Strengthening the Reporting of Observational Studies in Epidemiology (STROBE) guideline (**S4 Checklist**).

## Ethics Statement

This retrospective study used de-identified data obtained from the United Network for Organ Sharing (UNOS) registry. The study received an exemption from the George Mason University Institutional Review Board because it involved secondary analysis of de-identified registry data without access to information that could identify individual participants. The requirement for informed consent was waived.

### Study population and outcome definition

The study population included kidney transplant recipients with post-transplant prostate cancer, classified into two clinically distinct phenotypes. De novo prostate cancer was defined as prostate cancer diagnosed after transplantation in recipients without a documented history of prostate malignancy prior to transplant. Recurrent prostate cancer was defined as recurrence after transplantation in recipients with a known history of prostate cancer before transplant. Phenotype classification was based on registry indicators together with corresponding diagnosis dates. No patients met criteria for both categories, confirming mutually exclusive classification. The primary outcome was defined as a binary variable indicating recurrent prostate cancer compared with de novo prostate cancer.

The primary cohort included 660 recipients, of whom 623 had de novo disease and 37 had recurrent disease. A secondary cohort restricted to deceased donor recipients included 468 patients, excluding 192 living donor recipients in whom donor variables such as kidney donor profile index (KDPI), donor creatinine, and expanded criteria donor (ECD) status were not defined. These cohorts were used for complementary analyses, with the full cohort used for primary analyses and the deceased donor cohort used for analyses involving donor variables.

Diagnosis date variables were used to verify phenotype classification and to describe timing of cancer occurrence but were not included as predictors in the primary models. The registry does not distinguish between biochemical, local, or metastatic recurrence, and these were therefore analyzed as a single outcome.

### Covariate selection

Candidate covariates were selected a priori based on clinical relevance and prior work in transplant oncology. Variables were grouped into recipient, donor, and transplant, immunologic, and treatment domains. Recipient characteristics included age, body mass index (BMI), functional status, dialysis status at transplant, calculated panel reactive antibody (cPRA), retransplantation status, and serum creatinine. Donor and transplant factors included donor age, donor creatinine, donation after circulatory death (DCD), ECD status, allocation geography, and transplant year. Donor organ quality was further represented using the KDPI in analyses restricted to deceased donor recipients.

Immunologic variables included human leukocyte antigen (HLA) mismatch and serum albumin. Immunosuppression exposure was defined using regimen-specific indicators. Induction therapy was categorized as anti-thymocyte globulin (ATG), interleukin-2 receptor (IL-2R) antagonist, alemtuzumab, ATG plus an IL-2R antagonist, or no induction. ATG served as the reference category. Maintenance therapy was defined using calcineurin inhibitor (CNI) and mycophenolate mofetil (MMF) regimens with or without steroids, with the steroid-free regimen serving as the reference. Regimens containing mammalian target of rapamycin (mTOR) inhibitors were not evaluated because no recipients in the analytic cohort received these therapies.

## Statistical analysis

### Data preprocessing and missing data handling

All variables were reviewed for consistency prior to analysis. Binary variables were coded as indicator variables; continuous variables were retained in numeric form, and categorical variables were harmonized. Functional status was summarized using detailed Karnofsky Performance Score (KPS) categories for descriptive analyses (80–100, 70, 50–60, 10–40, and unknown) and collapsed into broader groups (KPS ≥70, 50–60, ≤40, and unknown) for modeling to reduce sparse cells. KPS ≥70 served as the reference category. Missing data were assessed for frequency and pattern. Donor variables such as KDPI, donor creatinine, and ECD status were structurally undefined in living donor transplants and were therefore not imputed. These variables were included only in analyses restricted to the deceased donor cohort.

Variables with very high missingness (greater than 80 percent) were excluded from analysis to avoid unstable estimates. Variables representing post-transplant events, including early rejection, were not included in primary models because of concerns regarding temporal ordering. For variables with low levels of missingness (less than 5 percent), missing values were imputed using median values for continuous variables and mode values for categorical variables. Given the limited degree of missingness, this approach was used to preserve sample size without materially affecting estimates.

### Descriptive analysis

Baseline characteristics were summarized using medians and interquartile ranges for continuous variables and counts and percentages for categorical variables. Differences between groups were assessed using the Mann Whitney U test for continuous variables and chi square or Fisher exact tests for categorical variables as appropriate. Standardized mean differences were calculated to assess the magnitude of differences between groups, with an absolute value greater than 0.10 considered indicative of meaningful imbalance. Transplant year was categorized into clinically meaningful eras to account for temporal changes in transplant practice and immunosuppression strategies.

### Multivariable analysis

Factors associated with prostate cancer phenotype were evaluated using logistic regression. Given the small number of recurrent events (n = 37), Firth penalized logistic regression was used to reduce small sample bias and to address potential separation. Standard logistic regression can produce unstable estimates in this setting; Firth regression provides more reliable estimates by penalizing the likelihood [29].

The primary model was restricted to a small number of prespecified clinically relevant variables to limit overfitting and ensure an appropriate ratio of predictors to outcome events, consistent with established guidance for regression modeling in studies with limited outcome events [30,31]. Variables were selected based on clinical relevance and consistency with prior literature rather than automated selection procedures. Model calibration was assessed using calibration plots and the Brier score by comparing predicted probabilities of recurrent phenotype with observed outcomes. Multicollinearity among covariates was evaluated using variance inflation factors, with values greater than 5 considered indicative of potential collinearity.

### Sensitivity analyses

Several sensitivity analyses were conducted to assess the robustness of the findings. These included models restricted to deceased donor recipients, models incorporating donor organ quality variables, and reduced variable models. An additional model incorporated time from transplantation to prostate cancer diagnosis to evaluate whether timing influenced the observed associations. The consistency of findings across these analyses was used to support the validity of the primary results.

### Machine learning analysis

To complement the regression analyses and explore potential nonlinear relationships, supervised machine learning models were developed to classify prostate cancer phenotype. Models included elastic net logistic regression, random forest, and extreme gradient boosting (XGBoost). Model development used a nested cross-validation framework to limit overfitting. The outer loop consisted of fivefold stratified cross-validation for performance estimation, while hyperparameters were tuned within an inner threefold stratified cross-validation loop. Final model performance was evaluated using out-of-fold predictions from the outer loop.

Model performance was assessed using the area under the receiver operating characteristic curve (ROC AUC), precision recall area under the curve (PR-AUC), and the Brier score. Calibration was evaluated using calibration plots and by estimating calibration intercept and slope. Class imbalance was addressed using class weighting for logistic and tree models and scale adjustment for gradient boosting. Model interpretability was assessed using feature importance for tree models and Shapley value analysis for the gradient boosting model. Given the limited number of recurrent events, machine learning analyses were pre-specified as exploratory. These analyses were used to assess whether patterns observed in the regression models were consistent across alternative modeling approaches rather than to develop definitive prediction models.

## Statistical software

All analyses were conducted in Python using pandas, NumPy, SciPy, statsmodels, scikit-learn, and XGBoost. Firth logistic regression was implemented using the logistf package in R via the rpy2 interface. Statistical significance was defined using two-sided tests with a significance level of 0.05.

## Results

### Cohort characteristics

The study cohort included 660 kidney transplant recipients with post-transplant prostate cancer, of whom 623 (94.4%) had de novo prostate cancer and 37 (5.6%) had recurrent prostate cancer. The median recipient age was 63 years (IQR 57.8–68.0) overall, with recipients in the recurrent group older than those in the de novo group [67 years (IQR 64–72) vs 63 years (IQR 57–67), p < 0.001; SMD 0.802]. The distribution of recipient age by phenotype is shown in **Fig 1**.

**Fig 1.**
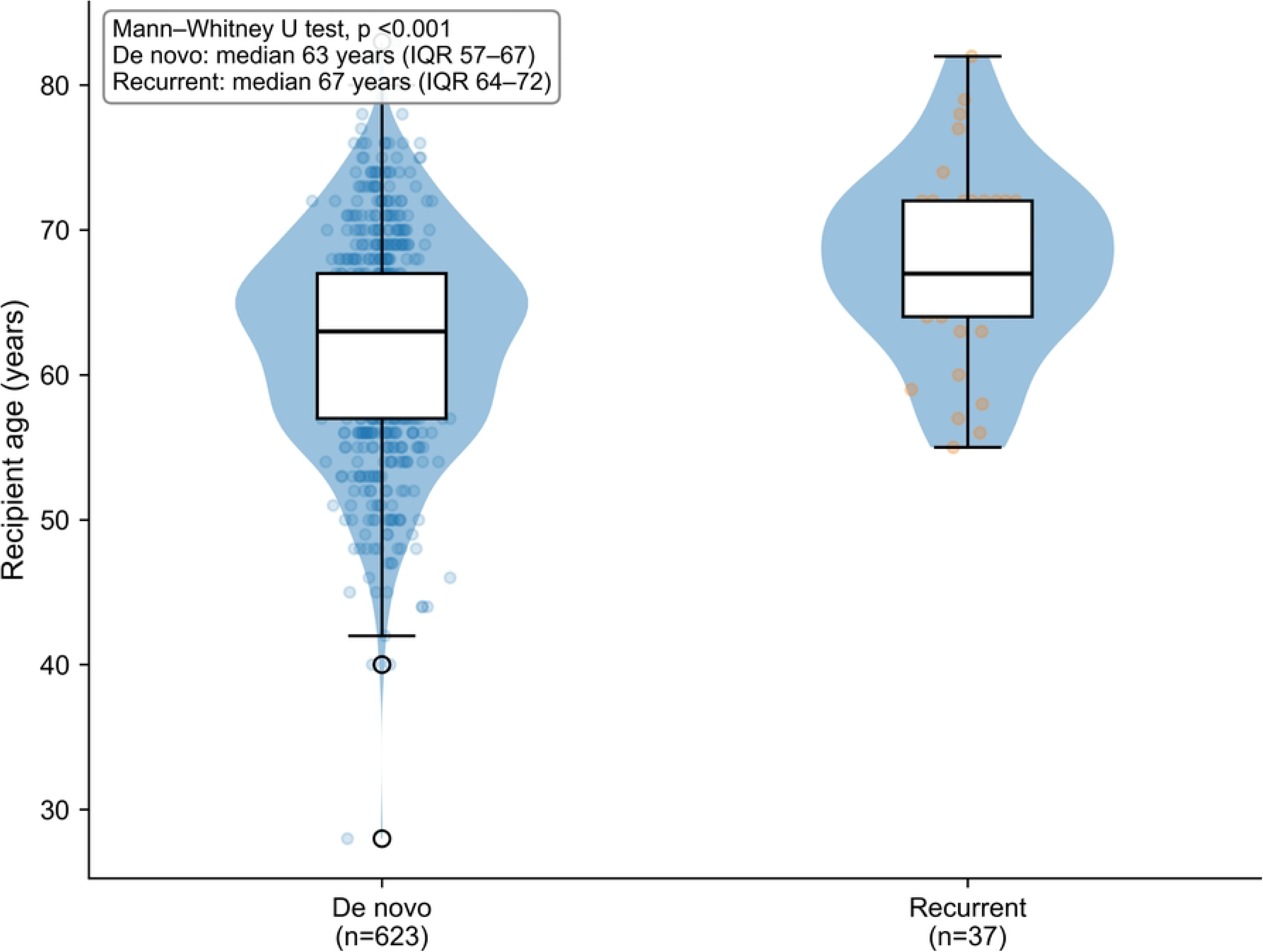
Recipient age by prostate cancer phenotype after kidney transplantation. Violin and box plots show the distribution of recipient age among recipients with de novo and recurrent prostate cancer. Boxes represent the interquartile range; horizontal lines indicate medians, and violin width reflects the distribution of the data.

The racial and ethnic composition of the cohort included 278 White recipients (42.1%), 282 Black recipients (42.7%), 74 Hispanic recipients (11.2%), and 23 Asian recipients (3.5%). Compared with the de novo group, the recurrent group had a slightly higher proportion of White recipients (48.6% vs 41.7%) and Hispanic recipients (18.9% vs 10.8%), and a lower proportion of Black recipients (32.4% vs 43.3%). No Asian recipients were observed in the recurrent group. Recipient BMI was lower in the recurrent group [26.5 (IQR 25.1–29.7) vs 28.3 (IQR 25.4–31.7), p = 0.028; SMD 0.402]. Diabetes was present in 273 recipients (41.4%) and was similar between groups. Dialysis before transplantation was common, occurring in 486 recipients (73.6%). Fifty recipients (7.6%) underwent kidney retransplantation, whereas none of the recipients with recurrent prostate cancer had undergone retransplantation.

Functional status was generally preserved, with 53.6% of recipients classified as independent or near independent and 25.5% able to perform self-care but unable to work. Lower functional status categories were uncommon (1.8%), and the overall distribution did not differ between groups (p = 0.467). Serum albumin was similar between groups [4.0 (IQR 3.7–4.3) vs 4.1 (IQR 3.7–4.3)]. Overall cPRA levels were low [median 0.0% (IQR 0.0–4.5)] but showed a moderate imbalance between groups (p = 0.111; SMD 0.292). HLA mismatch was similar across groups [median 4.0 (IQR 3.0–5.0)].

Deceased donor transplants accounted for 468 cases (70.9%) and living donor transplants for 192 cases (29.1%). The proportion of deceased donor transplantation was similar between de novo and recurrent phenotypes (71.1% vs 67.6%). The median donor age was 46 years (IQR 33.8–55.0) overall and was higher in the recurrent group than in the de novo group [53 years (IQR 44–58) vs 46 years (IQR 33–55), p = 0.008; SMD 0.490]. Donor diabetes, donor hypertension, donor BMI, donor creatinine, DCD, ECD status, and cold ischemia time were broadly similar between groups. Among donor organ quality measures, KDPI was 0.4 (IQR 0.2–0.6) overall and did not differ meaningfully between groups.

Most recipients received ATG induction (56.2%), followed by IL-2R antagonist induction (24.4%), alemtuzumab (12.3%), and no induction (5.6%). ATG + IL-2R antagonist induction was uncommon and observed in 10 recipients (1.5%). IL-2R antagonist induction was more frequent among recipients with recurrent prostate cancer than among those with de novo disease (40.5% vs 23.4%, p = 0.031; SMD 0.367), whereas ATG induction was less frequent in the recurrent group (45.9% vs 56.8%). For maintenance therapy, 476 recipients (72.1%) received CNI + MMF + steroids and 184 (27.9%) received CNI + MMF without steroids. No recipients in the analytic cohort received maintenance regimens containing mTOR inhibitors. Full baseline characteristics are presented in **Table 1**.

**Table 1.** Baseline characteristics of kidney transplant recipients with post-transplant prostate cancer, by phenotype.

| Characteristic | Overall (N=660) | De novo (n=623) | Recurrent (n=37) | P-value | SMD |
| --- | --- | --- | --- | --- | --- |
| **Recipient characteristics** |  |  |  |  |  |
| Age, years, median (IQR) | 63.0 (57.8–68.0) | 63.0 (57.0–67.0) | 67.0 (64.0–72.0) | <0.001 | 0.802 |
| White race, n (%) | 278 (42.1) | 260 (41.7) | 18 (48.6) | 0.512 | 0.139 |
| Black race, n (%) | 282 (42.7) | 270 (43.3) | 12 (32.4) | 0.258 | 0.225 |
| Hispanic ethnicity, n (%) | 74 (11.2) | 67 (10.8) | 7 (18.9) | 0.207 | 0.230 |
| Asian race, n (%) | 23 (3.5) | 23 (3.7) | 0 (0.0) | 0.633 | 0.274 |
| BMI, kg/m<sup>2</sup>, median (IQR) | 28.2 (25.3–31.5) | 28.3 (25.4–31.7) | 26.5 (25.1–29.7) | 0.028 | 0.402 |
| Diabetes, n (%) | 273 (41.4) | 260 (41.7) | 13 (35.1) | 0.535 | 0.136 |
| Peripheral vascular disease, n (%) | 80 (12.1) | 74 (11.9) | 6 (16.2) | 0.599 | 0.125 |
| Functional status, n (%) | – | – | – | 0.467 | – |
| 80–100: Independent or near independent | 354 (53.6) | 330 (53.0) | 24 (64.9) | – | 0.242 |
| 70: Self-care, unable to work normally | 168 (25.5) | 159 (25.5) | 9 (24.3) | – | 0.028 |
| 50–60: Requires assistance | 104 (15.8) | 100 (16.1) | 4 (10.8) | – | 0.154 |
| 10–40: Disabled or severely impaired | 12 (1.8) | 12 (1.9) | 0 (0.0) | – | 0.197 |
| Unknown | 22 (3.3) | 22 (3.5) | 0 (0.0) | – | 0.268 |
| Serum albumin, g/dL, median (IQR) | 4.0 (3.7–4.3) | 4.0 (3.7–4.3) | 4.1 (3.7–4.3) | 0.754 | 0.078 |
| cPRA, %, median (IQR) | 0.0 (0.0–4.5) | 0.0 (0.0–4.5) | 0.0 (0.0–0.0) | 0.111 | 0.292 |
| Retransplantation, n (%) | 50 (7.6) | 50 (8.0) | 0 (0.0) | 0.102 | 0.409 |
| On dialysis at transplant, n (%) | 486 (73.6) | 457 (73.4) | 29 (78.4) | 0.63 | 0.117 |
| Wait time, days, median (IQR) | 463.5 (143.5–1125.5) | 460.0 (140.5–1128.0) | 617.0 (176.0–1118.0) | 0.863 | 0.014 |
| Recipient creatinine, mg/dL, median (IQR) | 7.9 (5.4–10.5) | 7.9 (5.4–10.5) | 7.3 (5.3–9.1) | 0.448 | 0.159 |
| **Donor and transplant characteristics** |  |  |  |  |  |
| Donor age, years, median (IQR) | 46.0 (33.8–55.0) | 46.0 (33.0–55.0) | 53.0 (44.0–58.0) | 0.008 | 0.49 |
| Male donor, n (%) | 359 (54.4) | 340 (54.6) | 19 (51.4) | 0.832 | 0.065 |
| White race, n (%) | 474 (71.8) | 449 (72.1) | 25 (67.6) | 0.687 | 0.098 |
| Black race, n (%) | 75 (11.4) | 72 (11.6) | 3 (8.1) | 0.789 | 0.116 |
| Hispanic ethnicity, n (%) | 88 (13.3) | 81 (13.0) | 7 (18.9) | 0.435 | 0.162 |
| Asian race, n (%) | 17 (2.6) | 15 (2.4) | 2 (5.4) | 0.246 | 0.155 |
| Diabetes, n (%) | 33 (5.0) | 31 (5.0) | 2 (5.4) | 0.707 | 0.019 |
| Donor hypertension, n (%) | 163 (24.7) | 153 (24.6) | 10 (27.0) | 0.887 | 0.056 |
| Donor BMI, kg/m<sup>2</sup>, median (IQR) | 27.0 (23.9–30.9) | 27.0 (23.8–30.8) | 27.3 (24.4–32.0) | 0.415 | 0.029 |
| Donor type, n (%) | – | – | – | 0.784 | 0.077 |
| Deceased | 468 (70.9) | 443 (71.1) | 25 (67.6) | – | – |
| Living | 192 (29.1) | 180 (28.9) | 12 (32.4) | – | – |
| Donor creatinine, mg/dL, median (IQR) | 0.9 (0.7–1.3) | 0.9 (0.7–1.3) | 1.0 (0.8–1.1) | 0.555 | 0.008 |
| KDPI, median (IQR) | 0.4 (0.2–0.6) | 0.4 (0.2–0.6) | 0.5 (0.3–0.6) | 0.484 | 0.115 |
| Donation after circulatory death, n (%) | 114 (17.3) | 108 (17.3) | 6 (16.2) | 1 | 0.03 |
| Expanded criteria donor, n (%) | 83 (17.7) | 78 (17.6) | 5 (20.0) | 0.972 | 0.061 |
| Cold ischemia time, hours, median (IQR) | 14.3 (4.7–21.0) | 14.3 (4.8–21.2) | 14.4 (1.5–19.7) | 0.669 | 0.017 |
| Allocation, n (%) | – | – | – | – | – |
| Local | 489 (74.1) | 462 (74.2) | 27 (73.0) | 1 | 0.027 |
| Regional | 80 (12.1) | 74 (11.9) | 6 (16.2) | 0.599 | 0.125 |
| National | 91 (13.8) | 87 (14.0) | 4 (10.8) | 0.806 | 0.096 |
| Transplant era, n (%) | – | – | – | 0.221 | 0.229 |
| 2015–2019 | 477 (72.3) | 454 (72.9) | 23 (62.2) | – | – |
| 2020–2024 | 183 (27.7) | 169 (27.1) | 14 (37.8) | – | – |
| **Immunosuppression** |  |  |  |  |  |
| Induction therapy, n (%) | – | – | – | – | – |
| ATG | 371 (56.2) | 354 (56.8) | 17 (45.9) | 0.261 | 0.218 |
| IL-2R antagonist | 161 (24.4) | 146 (23.4) | 15 (40.5) | 0.031 | 0.367 |
| Alemtuzumab | 81 (12.3) | 76 (12.2) | 5 (13.5) | 1 | 0.039 |
| ATG + IL-2R antagonist | 10 (1.5%) | 10 (1.6%) | 0 (0.0%) | 1.000 | 0.100 |
| None | 37 (5.6) | 37 (5.9) | 0 (0.0) | 0.257 | 0.35 |
| Maintenance therapy, n (%) | – | – | – | – | – |
| CNI + MMF | 184 (27.9) | 176 (28.3) | 8 (21.6) | 0.493 | 0.153 |
| CNI + MMF + steroids | 476 (72.1) | 447 (71.7) | 29 (78.4) | – | 0.153 |
| **Immunologic variables** |  |  |  |  |  |
| HLA mismatch, median (IQR) | 4.0 (3.0–5.0) | 4.0 (3.0–5.0) | 4.0 (4.0–5.0) | 0.893 | 0.121 |

Continuous variables are summarized as median (IQR), and categorical variables as number (%). Between-group comparisons were performed using the Mann–Whitney U test for continuous variables and the chi-square test or Fisher's exact test for categorical variables, as appropriate. For grouped categorical variables, p values are reported at the overall category level rather than for individual subcategories. "–" indicates that subgroup p values were not calculated separately. Standardized mean differences (SMDs) are reported as absolute values; an SMD greater than 0.10 was considered indicative of meaningful imbalance. Functional status was derived from Karnofsky Performance Score categories. Donor creatinine, KDPI, and expanded criteria donor status are defined only for deceased donor transplants and should therefore be interpreted within the deceased donor subset. No recipients received mTOR inhibitor maintenance regimens in this analytic cohort. All recipients in the cohort were male. ATG, anti-thymocyte globulin; CNI, calcineurin inhibitor; cPRA, calculated panel reactive antibody; HLA, human leukocyte antigen; IL-2R, interleukin 2 receptor; KDPI, kidney donor profile index; MMF, mycophenolate mofetil; SMD, standardized mean difference.

Time from kidney transplantation to prostate cancer diagnosis was examined. The median time to diagnosis was 2.61 years (IQR 1.33–4.38) among de novo cases and 2.21 years (IQR 0.84– 4.11) among recurrent cases, with no statistically significant difference between groups (p = 0.202).

The distribution of time from transplant to diagnosis by phenotype is shown in **S1 Fig.**

### Multivariable analysis

In the primary Firth penalized logistic regression model, recipient age was independently associated with recurrent versus de novo prostate cancer phenotype. The model included a limited number of prespecified variables relative to the number of recurrent events (n = 37) to reduce the risk of overfitting. Each one-year increase in recipient age was associated with higher odds of recurrent disease (adjusted odds ratio [OR] 1.11, 95% CI 1.05–1.17, p < 0.001). Adjusted estimates for all covariates are presented in **Table 2** and illustrated in **Fig 2**.

**Fig 2.**
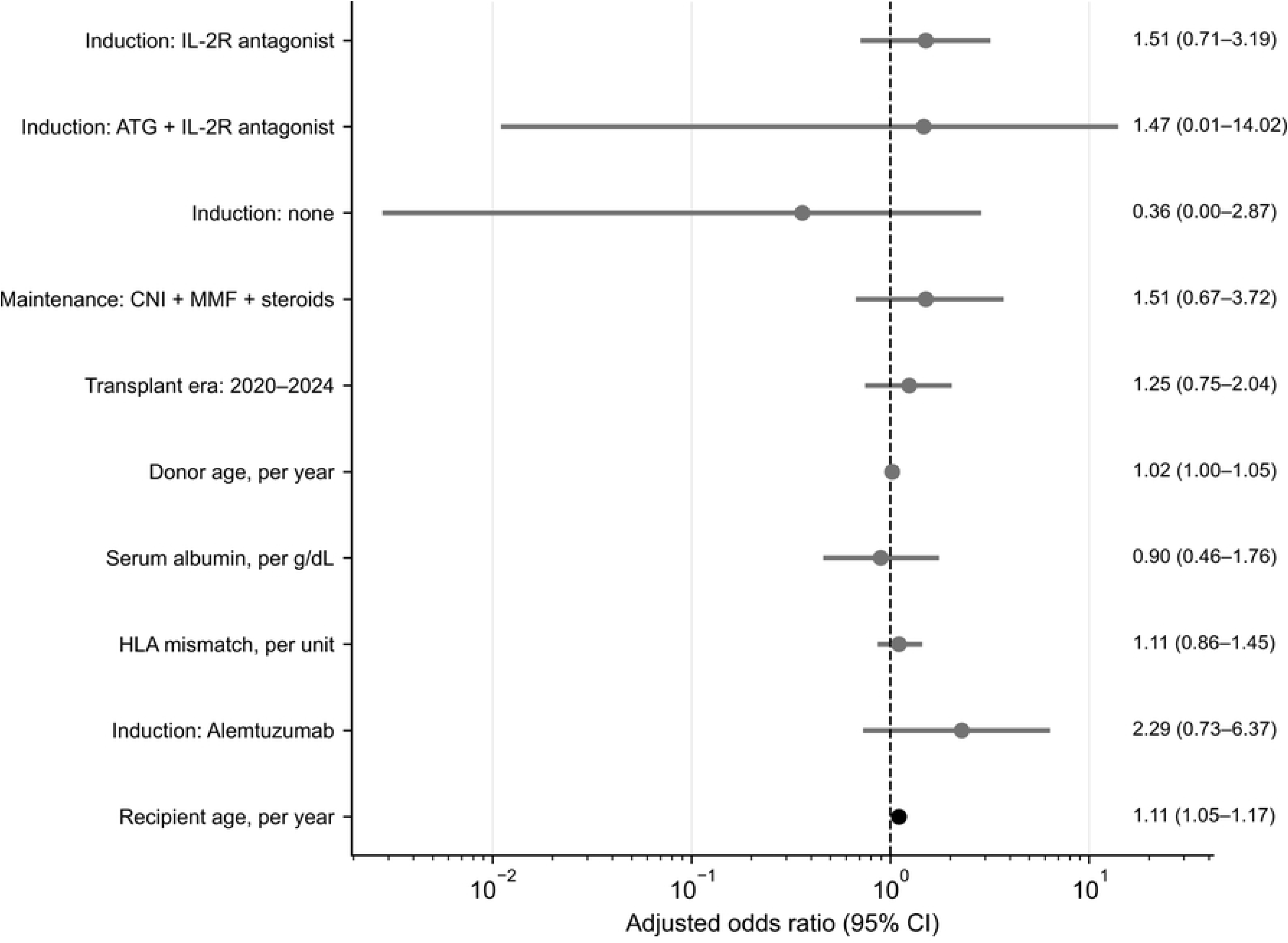
Forest plot showing adjusted odds ratios from the primary Firth penalized logistic regression model. Each marker corresponds to an adjusted odds ratio, and the accompanying horizontal bar indicates its 95% confidence interval. The vertical dashed line marks an odds ratio of 1.0. Estimates to the right of the reference line indicate higher odds of recurrent compared with de novo prostate cancer phenotype. The statistically significant association (p < 0.05) is indicated by a filled square, whereas non-significant estimates are shown as filled circles. Reference categories were anti-thymocyte globulin (ATG) for induction therapy, calcineurin inhibitor plus mycophenolate mofetil without steroids (CNI + MMF) for maintenance therapy, and transplant era 2015–2019 for transplant era comparisons. CI, confidence interval; HLA, human leukocyte antigen; IL-2R, interleukin-2 receptor; MMF, mycophenolate mofetil; OR, odds ratio.

**Table 2.** Primary Firth penalized logistic regression model for recurrent versus de novo prostate cancer.

| Covariate | Adjusted OR (95% CI) | P-value |
| --- | --- | --- |
| Recipient age, per year | 1.11 (1.05–1.17) | <0.001 |
| Induction therapy |  |  |
| ATG | Reference | – |
| IL-2R antagonist | 1.51 (0.71–3.19) | 0.283 |
| Alemtuzumab | 2.29 (0.73–6.37) | 0.148 |
| None | 0.36 (0.00–2.87) | 0.413 |
| ATG + IL-2R antagonist | 1.47 (0.01–14.02) | 0.806 |
| Maintenance therapy |  |  |
| CNI + MMF | Reference | – |
| CNI + MMF + steroids | 1.51 (0.67–3.72) | 0.331 |
| HLA mismatch, per unit | 1.11 (0.86–1.45) | 0.440 |
| Serum albumin, per g/dL | 0.90 (0.46–1.76) | 0.749 |
| Donor age, per year | 1.02 (1.00–1.05) | 0.100 |
| Transplant era, 2020–2024 vs 2015–2019 | 1.25 (0.75–2.04) | 0.389 |

Odds ratios were estimated using Firth penalized logistic regression to reduce small sample bias and address sparse outcome events. The outcome was recurrent prostate cancer compared with de novo prostate cancer. ATG was the reference category for induction therapy, CNI + MMF for maintenance therapy, and 2015–2019 for transplant era. No recipients received mTOR inhibitor maintenance regimens in this cohort; these exposures were not evaluated. The “None” and “ATG + IL-2R antagonist” induction categories included zero or very few recurrent events; confidence intervals are correspondingly wide, and these estimates should be interpreted with caution. ATG indicates anti-thymocyte globulin; CI, confidence intervals; CNI, calcineurin inhibitor; HLA, human leukocyte antigen; IL-2R, interleukin 2 receptor; MMF, mycophenolate mofetil; OR, odds ratio.

No other covariates were significantly associated with prostate cancer phenotype in the primary model (**Table 2**). Induction with IL-2R antagonist (OR 1.51, 95% CI 0.71–3.19, p = 0.28) and alemtuzumab (OR 2.29, 95% CI 0.73–6.37, p = 0.15) were not significantly associated with recurrent phenotype compared with ATG. Maintenance therapy with CNI + MMF + steroids was similarly not associated with phenotype (OR 1.51, 95% CI 0.67–3.72, p = 0.33).

Immunologic and clinical variables, including HLA mismatch (OR 1.11, 95% CI 0.86–1.45, p = 0.44), serum albumin (OR 0.90, 95% CI 0.46–1.76, p = 0.75), and transplant era (OR 1.25, 95% CI 0.75–2.04, p = 0.39), were not associated with prostate cancer phenotype. Donor age showed a small increase in odds of recurrent phenotype that did not reach statistical significance (OR 1.02, 95% CI 1.00–1.05, p = 0.10).

Variance inflation factors for all covariates were below 5, indicating no evidence of problematic multicollinearity. Model calibration for the primary Firth logistic regression is shown in **Fig 3**. Predicted probabilities were well aligned with observed event rates across risk strata. The Brier score for the primary model was 0.050, compared with a null model Brier score of 0.053 based on the observed event rate, corresponding to modest improvement in overall probabilistic accuracy.

**Fig 3.**
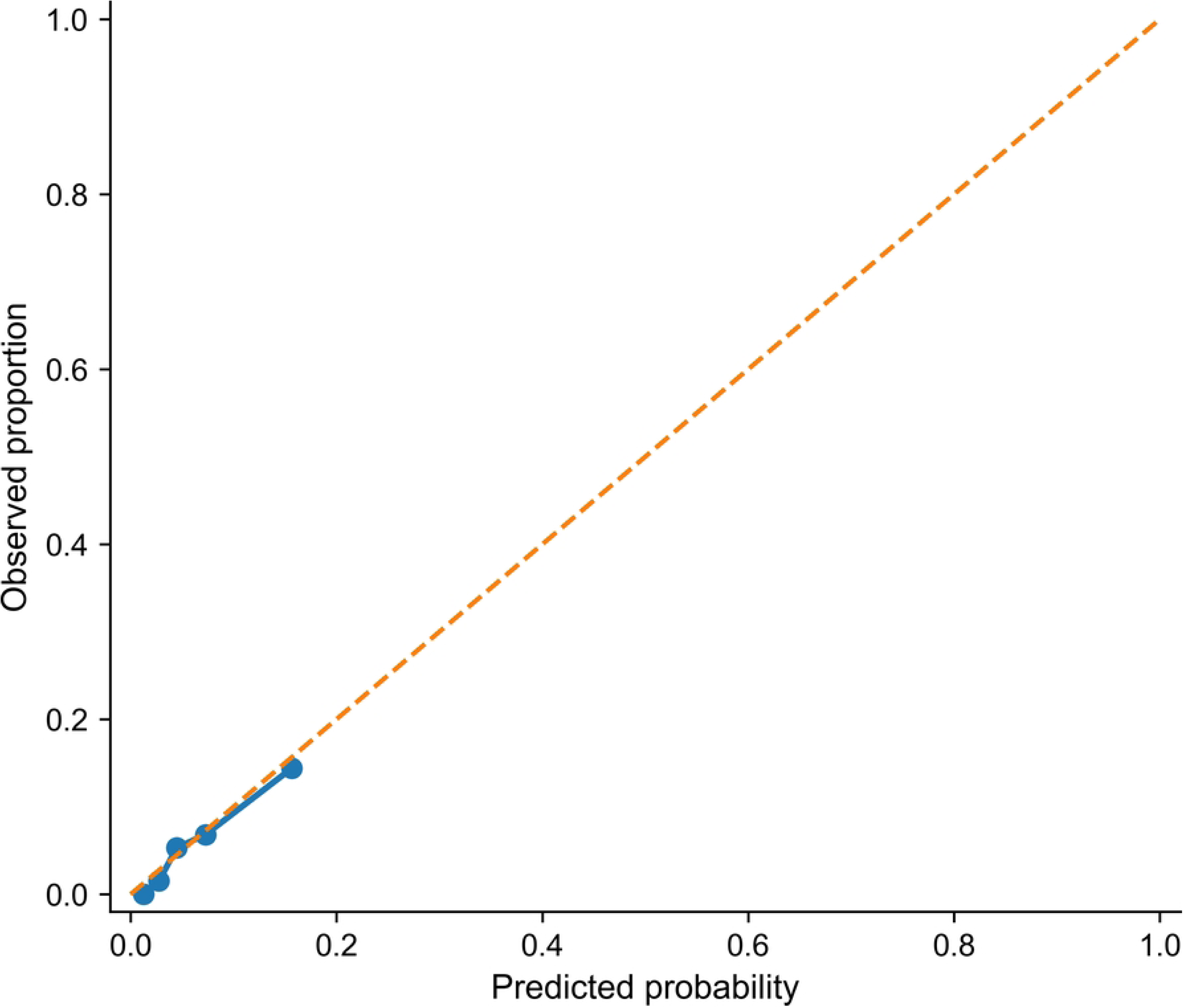
Calibration of the primary Firth logistic regression model. Calibration plot comparing predicted and observed probabilities of recurrent prostate cancer phenotype across risk groups. The dashed line represents perfect calibration.

### Sensitivity analyses

The association between recipient age and recurrent prostate cancer phenotype was consistent across all sensitivity analyses. In models additionally adjusted for functional status, the association with recipient age was unchanged (OR 1.11, 95% CI 1.05–1.18, p < 0.001). Functional status categories were not independently associated with phenotype, and CIs were wide for lower functional status categories, reflecting sparse event counts in these groups.

Findings were unchanged after additional adjustment for dialysis status and retransplantation. The association with recipient age remained consistent (OR 1.10, 95% CI 1.05–1.17, p < 0.001). Dialysis at transplant (OR 1.52, 95% CI 0.69–3.67, p = 0.31) and retransplantation (OR 0.27, 95% CI 0.00–2.07, p = 0.26) were not associated with phenotype; no recurrent cases were observed among retransplant recipients, resulting in wide CIs and limited interpretability.

In analyses restricted to deceased donor recipients, the association with recipient age remained consistent (OR 1.14, 95% CI 1.06–1.23, p < 0.001). In this cohort, alemtuzumab induction was associated with higher odds of recurrent phenotype compared with ATG (OR 4.92, 95% CI 1.25–17.79, p = 0.024), although this finding was based on a small number of events. Maintenance therapy with CNI + MMF + steroids demonstrated a higher point estimate compared with CNI + MMF without steroids, although the confidence interval included the null (OR 3.13, 95% CI 0.97–13.66, p = 0.057). Donor and transplant variables, including donor creatinine, DCD, ECD status, recipient creatinine, and transplant era, were not associated with phenotype.

In a model using KDPI as a summary measure of donor organ quality, the association with recipient age remained consistent (OR 1.14, 95% CI 1.06–1.23, p < 0.001). Alemtuzumab induction remained associated with recurrent phenotype (OR 4.13, 95% CI 1.10–13.86, p = 0.037), and maintenance therapy with CNI + MMF + steroids was associated with recurrent phenotype (OR 3.31, 95% CI 1.05–13.85, p = 0.040). KDPI was not associated with phenotype, with wide CIs reflecting limited precision (OR 0.57, 95% CI 0.09–3.33, p = 0.53).

In models incorporating time from transplantation to prostate cancer diagnosis, time to diagnosis was not associated with phenotype (OR 1.06 per year, 95% CI 0.86–1.30, p = 0.58), and the association with recipient age was unchanged (OR 1.11, 95% CI 1.05–1.17, p < 0.001). Similar findings were observed in a deceased donor restricted timing model, in which time to diagnosis was not associated with phenotype (OR 1.16, 95% CI 0.95–1.41, p = 0.15). Detailed results for all sensitivity models are presented in **S1 Table**.

### Machine learning analysis

Exploratory machine learning analyses were conducted to complement the primary regression findings and to assess whether nonlinear methods improved classification of prostate cancer phenotype. Overall model performance was modest and comparable across approaches (**Table 3**). ROC AUC values were similar across models: 0.693 for elastic net logistic regression, 0.696 for random forest, and 0.700 for XGBoost, with overlapping confidence intervals across all models.

**Table 3.**
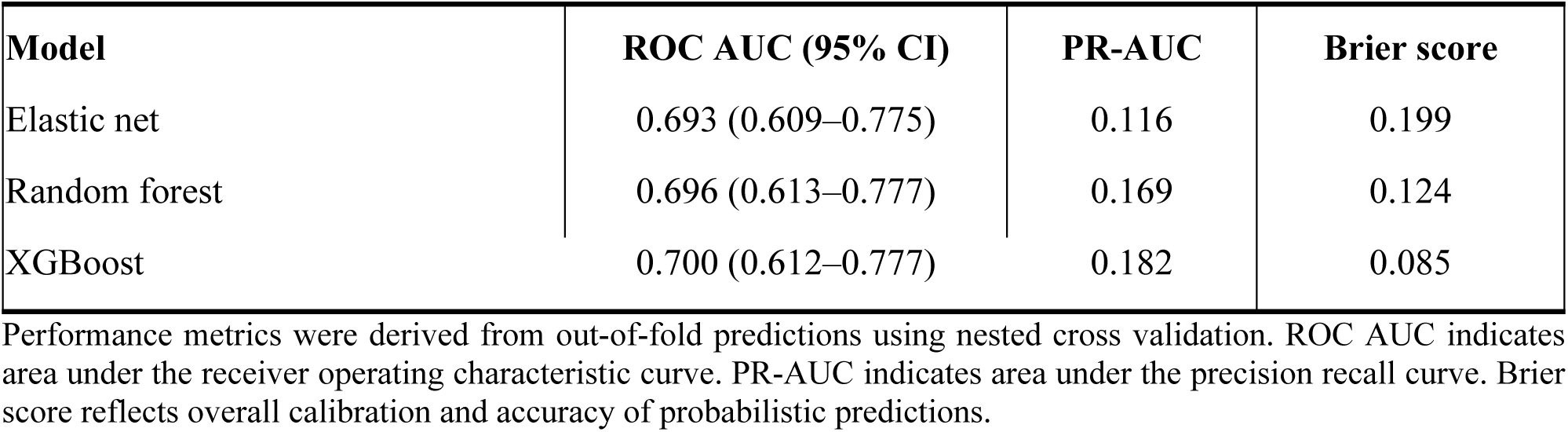
Performance of machine learning models for classification of prostate cancer phenotype.

Precision recall performance was limited. PR-AUC values were 0.116 for elastic net, 0.169 for random forest, and 0.182 for XGBoost, all above the baseline no-skill level corresponding to the prevalence of recurrent phenotype (PR-AUC = 0.056). Precision decreased at higher recall thresholds, consistent with class imbalance. Among the models, random forest demonstrated the highest PR-AUC, whereas XGBoost had the lowest Brier score. Discrimination performance is shown in **Fig 4**.

**Fig 4.**
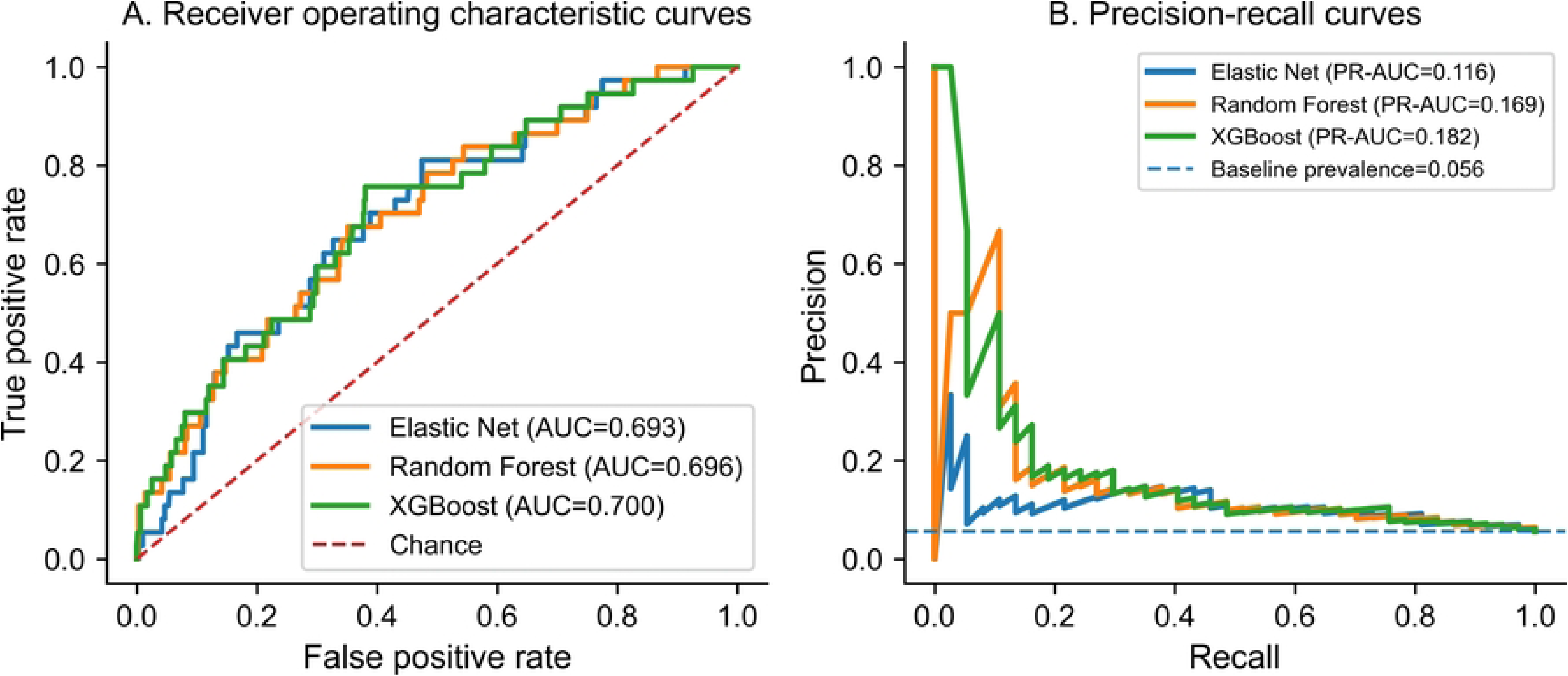
Discrimination performance of the machine learning models for classification of recurrent versus de novo prostate cancer phenotype. Receiver operating characteristic curves (panel A) and precision-recall curves (panel B) are shown for elastic net logistic regression, random forest, and XGBoost using out-of-fold predictions generated through nested cross-validation. The diagonal dashed line in panel A represents chance-level discrimination (AUC = 0.50). The dashed horizontal line in panel B indicates the baseline prevalence of recurrent prostate cancer in the study cohort (5.6%). Abbreviations: AUC, area under the receiver operating characteristic curve; PR-AUC, area under the precision-recall curve.

Calibration assessment showed systematic overestimation of recurrence risk across all models. Calibration intercepts were negative for elastic net, random forest, and XGBoost (indicating overestimation of predicted recurrence probabilities). Random forest had the calibration slope closest to 1, whereas XGBoost showed the lowest calibration slope. Calibration metrics are presented in **S2 Table**, and calibration plots are shown in **S2 Fig**.

Model interpretability analyses were consistent across approaches. In elastic net models, only recipient age and donor age retained nonzero coefficients after regularization. Random forest feature importance ranked recipient age, recipient BMI, and donor age as the most influential variables. In SHapley Additive exPlanations (SHAP) analysis of the XGBoost model, recipient age had the highest mean absolute SHAP value, followed by recipient BMI and donor age (**S3 Fig**.

Across the multivariable model and the machine learning analyses, recipient age was the most consistently identified variable. It was the only predictor retained after elastic net regularization, ranked highest in the random forest feature importance analysis, and demonstrated the largest mean absolute SHAP value in the XGBoost model. Given the limited number of recurrent events, these analyses were considered exploratory and were performed to determine whether similar patterns were observed across different analytical approaches rather than to develop a clinical prediction model.

## Discussion

In this national registry analysis of kidney transplant recipients who developed post-transplant prostate cancer, recipient age emerged as the only variable consistently associated with recurrent prostate cancer phenotype. This association remained stable across all prespecified sensitivity analyses, including adjustment for functional status, dialysis exposure, retransplantation, donor characteristics, and time from transplantation to cancer diagnosis. In contrast, immunologic factors, donor organ quality measures, and most immunosuppressive exposures were not independently associated with phenotype in the primary cohort. These findings suggest that recipients who develop recurrent prostate cancer after transplantation differ from those who develop de novo disease primarily on the basis of recipient characteristics rather than transplant exposures. More broadly, the results support the view that recurrent and de novo prostate cancer represent distinct clinical presentations among kidney transplant recipients who develop post-transplant prostate cancer.

### Recipient age as the primary determinant of prostate cancer phenotype

The central finding of this study was that each additional year of recipient age conferred an 11% increase in the odds of recurrent versus de novo prostate cancer phenotype (OR 1.11, 95% CI 1.05–1.17). The observed median age difference between recurrent and de novo cases (67 vs 63 years) is clinically meaningful. The prominence of age is consistent with the broader epidemiology of prostate cancer, where age is the strongest known risk factor for incidence, progression, and recurrence, likely reflecting cumulative biological and hormonal influences over time [16,32,33]. The persistence of this association despite chronic immunosuppression suggests that prostate cancer phenotype in transplant recipients may be influenced more by characteristics of the recipient and the cancer itself than by factors associated with transplantation. The modest elevation in donor age observed in the recurrent group did not retain independent significance in multivariable models. This likely reflects donor-recipient age concordance patterns in transplant allocation rather than a direct biological effect of donor age on prostate cancer phenotype. In the absence of a clear mechanistic pathway, this finding should be interpreted cautiously and more likely reflects allocation practices than a causal relationship.

### Absence of immunosuppression effects in the primary cohort

The absence of a significant association between immunosuppressive regimen and prostate cancer phenotype in the primary cohort is consistent with the broader evidence suggesting that prostate cancer is not strongly driven by immunosuppression. Unlike malignancies closely linked to impaired immune surveillance, such as post-transplant lymphoproliferative disorder, Kaposi sarcoma, and non-melanoma skin cancers, prostate cancer has not demonstrated a consistent increase in incidence among kidney transplant recipients in large registry studies and meta-analyses [19,20]. The current findings extend this observation by suggesting that immunosuppressive exposure may also have a limited role in determining prostate cancer phenotype after transplantation.

Large transplant registry studies have similarly shown that breast and prostate cancer do not exhibit the markedly elevated post-transplant incidence patterns observed in malignancies more directly related to immune dysfunction [7,8]. Both cancers are hormonally influenced malignancies and share important hereditary susceptibility pathways, including BRCA1 and BRCA2 associated risk [16]. The similarity in their epidemiologic behavior after transplantation suggests that hormonal influences and characteristics of the underlying disease may be more important than the degree of immunosuppression in shaping disease presentation. This interpretation is supported by prior work in breast cancer after transplantation, where post-transplant breast cancer has similarly been evaluated as a clinically meaningful entity with distinct outcomes [34].

### Sensitivity analyses in deceased donor recipients

Although immunosuppressive exposures were not significantly associated with prostate cancer phenotype in the primary cohort, several exploratory associations emerged in analyses restricted to deceased donor recipients. Alemtuzumab induction was associated with higher odds of recurrent phenotype compared with ATG in both the deceased donor model and the KDPI substitution model. In addition, maintenance therapy with CNI + MMF + steroids was associated with recurrent phenotype in the KDPI substitution model. These findings should be interpreted cautiously given the limited number of recurrent events, wide confidence intervals, and exploratory nature of the subgroup analyses.

If replicated in larger cohorts, several biologically plausible explanations may warrant consideration. Alemtuzumab produces profound and sustained lymphocyte depletion compared with other induction strategies, which could theoretically impair immune surveillance of residual malignant cells in recipients with a history of prostate cancer before transplantation [35,36]. Similarly, corticosteroid may also influence prostate cancer behavior through mechanisms that include modulation of androgen receptor activity and changes within the tumor microenvironment [37]. However, these observations should not be interpreted causally. In clinical practice, induction and maintenance regimens are not randomly assigned and are strongly influenced by recipient immunologic risk, sensitization status, retransplantation history, and center specific practice patterns. Residual confounding by indication therefore remains likely, particularly in analyses derived from national transplant registry data where detailed oncologic and immunologic variables are unavailable.

Importantly, donor characteristics, including donor creatinine, DCD, ECD status, and KDPI, were not independently associated with prostate cancer phenotype. Taken together, these findings suggest that any potential relationship between transplant exposures and phenotype is likely modest relative to the stronger influence of recipient level factors such as age.

### Timing of cancer diagnosis and phenotype classification

Time from transplantation to prostate cancer diagnosis was not independently associated with phenotype in either the primary model or the deceased donor restricted analyses. These findings suggest that differences in transplant to cancer interval do not explain the observed distribution of recurrent and de novo disease.

Although recipients with a prior history of prostate cancer may undergo closer oncologic follow-up after transplantation, recurrent cases were not identified significantly earlier after adjustment. The consistency of these findings across models suggests that recurrent and de novo prostate cancer may reflect clinically meaningful differences in disease presentation rather than being explained solely by the timing of post-transplant diagnosis.

### Machine learning findings and model performance

The machine learning analyses did not demonstrate improved discrimination compared with conventional multivariable logistic modeling. Model performance was modest across all methods, with similar ROC AUC values and overlapping confidence intervals for elastic net logistic regression, random forest, and XGBoost. Precision-recall performance was limited across models, consistent with the low prevalence of recurrent phenotype, although all models performed above the no skill baseline. Calibration assessment demonstrated systematic overestimation of recurrence risk across the machine learning models, whereas the primary Firth penalized logistic regression model showed closer agreement between predicted and observed probabilities. The magnitude of the negative calibration intercepts likely reflects the extreme class imbalance within the dataset, with recurrent phenotype accounting for only 5.6% of cases, together with the inherent difficulty of calibrating prediction models in the setting of sparse positive outcome events.

Across all analytic approaches, recipient age consistently emerged as the dominant predictor of prostate cancer phenotype. Recipient age was the only variable retained after elastic net regularization, ranked highest in random forest feature importance analyses, and demonstrated the largest mean absolute SHAP value in the XGBoost model. The consistency of these findings across the primary multivariable model and the machine learning analyses suggests that recipient age accounted for most of the information distinguishing recurrent from de novo prostate cancer in this cohort. More complex machine learning methods did not identify additional clinically meaningful patterns beyond those observed in the primary analysis.

These findings are consistent with prior methodological studies showing that machine learning models do not consistently outperform penalized regression approaches in clinical datasets with small sample sizes and low event rates, particularly when calibration and generalizability are considered [38,39]. The modest predictive performance observed across all models likely reflects the limited number of recurrent events and the absence of more granular oncologic variables, including tumor grade, prostate specific antigen kinetics, and treatment history, which are not captured within the registry dataset.

### Strengths

This study has several strengths. It is among the first analyses using national transplant registry data to examine differences between recurrent and de novo prostate cancer after kidney transplantation. By focusing on phenotype among recipients who developed post-transplant prostate cancer, the study provides insight into clinical, transplant, immunologic, and treatment characteristics associated with these distinct presentations. The use of a large transplant registry with detailed recipient, donor, transplant, and immunosuppressive variables enabled evaluation of multiple clinical domains within a real-world transplant population.

The analytic strategy was specifically designed to address methodological challenges associated with rare outcomes and sparse event data. Firth penalized logistic regression was used to reduce small sample bias and mitigate separation, and missing data handling incorporated explicit assessment of structural missingness in donor variables. Multiple prespecified sensitivity analyses were performed across different covariate structures and restricted cohorts, and the consistency of the primary findings across these models supports the robustness of the observed association between recipient age and prostate cancer phenotype.

An additional strength was the integration of complementary machine learning approaches as exploratory analyses. Although machine learning models did not improve predictive discrimination, the consistency of findings across multivariable regression and machine learning analyses provided additional support that recipient age was the strongest predictor of prostate cancer phenotype in this cohort.

### Limitations

The findings should be interpreted in light of several limitations. First, the number of recurrent prostate cancer events was small, which limited statistical power and constrained the number of covariates that could be included in multivariable models. Consequently, estimates for several variables, particularly immunosuppressive exposures, were imprecise and should be interpreted cautiously.

Second, the observational nature of the registry data precludes causal inference, and residual confounding remains possible, particularly for treatment related variables. Induction and maintenance immunosuppressive regimens are not randomly assigned in clinical practice and may reflect differences in recipient immunologic risk, comorbidity burden, transplant history, or center specific practice patterns that are incompletely captured within registry variables.

Third, the registry lacks detailed oncologic information, including tumor stage, Gleason grade, prostate specific antigen levels, treatment history, and evidence of biochemical recurrence before transplantation. The absence of these variables limits more granular characterization of prostate cancer phenotype and prevents assessment of disease severity at the time of transplant or recurrence.

Fourth, prostate cancer screening practices and surveillance intensity were not available within the registry and may have varied across transplant centers and study periods. Although timing analyses did not support surveillance interval alone as the primary explanation for phenotype differences, differential detection practices cannot be completely excluded.

Fifth, several donor characteristics, including donor creatinine, ECD status, and KDPI, were structurally unavailable for living donor transplants and therefore could only be evaluated in deceased donor restricted analyses.

Finally, this study reflects the national transplant experience captured within the registry during the study period and may not be fully generalizable to other healthcare systems or transplant populations with different immunosuppressive protocols or cancer surveillance practices.

## Conclusions

In this national registry cohort of kidney transplant recipients with post-transplant prostate cancer, recipient age was the most consistent factor associated with recurrent versus de novo prostate cancer. In contrast, immunosuppressive regimen, donor characteristics, and immunologic measures were not consistently associated with phenotype in the overall cohort. Overall, the observed differences between recurrent and de novo prostate cancer were more closely associated with recipient characteristics and the underlying disease than with the transplant factors examined in this cohort. This pattern differs from that observed in malignancies more closely linked to impaired immune surveillance and is consistent with the hypothesis that prostate cancer after transplantation may be influenced by mechanisms distinct from those driving many other post-transplant cancers. The consistency of the findings across the primary model, multiple sensitivity analyses, and exploratory machine learning analyses strengthens confidence in the observed association with age. Future studies with larger numbers of recurrent cases and more detailed oncologic information are needed to better understand the factors that distinguish recurrent from de novo prostate cancer after kidney transplantation.

## Acknowledgments

The authors thank the United Network for Organ Sharing (UNOS) for providing access to the de-identified registry data used in this study. The interpretations and conclusions presented are those of the authors and do not necessarily represent the views of UNOS or its affiliated organizations.

## Author Contributions

Conceptualization: Kunle Apanisile, Naoru Koizumi

Methodology: Kunle Apanisile, Meng-Hao Li, Naoru Koizumi

Formal analysis: Kunle Apanisile, Meng-Hao Li

Writing – original draft: Kunle Apanisile

Writing – review & editing: Naoru Koizumi, Giovanni Faddoul, Obi Ekwenna

Supervision: Naoru Koizumi, Giovanni Faddoul, Meng-Hao Li, Obi Ekwenna

## Funding

The authors received no specific funding for this work.

## Competing Interests

The authors declare that they have no competing interests.

## Data Availability

The data underlying this study are third-party data obtained from the Organ Procurement and Transplantation Network (OPTN), which is operated by the United Network for Organ Sharing (UNOS). These registry data contain sensitive, de-identified patient-level information and are subject to legal and contractual restrictions that prevent the authors from sharing them publicly. Qualified researchers may request access to the same dataset directly through the OPTN/UNOS data request process (https://unos.org/data/). Access is granted by UNOS following submission and approval of a formal data request and execution of a data use agreement. The authors did not receive any special access privileges unavailable to other researchers.

The analytical code used to construct the prostate cancer analytic cohort, perform data preprocessing, generate study variables, conduct statistical and machine learning analyses, and produce the tables and figures is publicly available at: https://github.com/Olukhunlay-hub/kidney-transplant-prostate-cancer-phenotype.

Researchers with access to the OPTN/UNOS registry can use the repository to reproduce the analytic cohort and all analyses presented in this manuscript.

## Supporting information

S1 Table. Sensitivity analyses using Firth penalized logistic regression.

S2 Table. Calibration metrics for machine learning models.

---

**S1 Fig.**
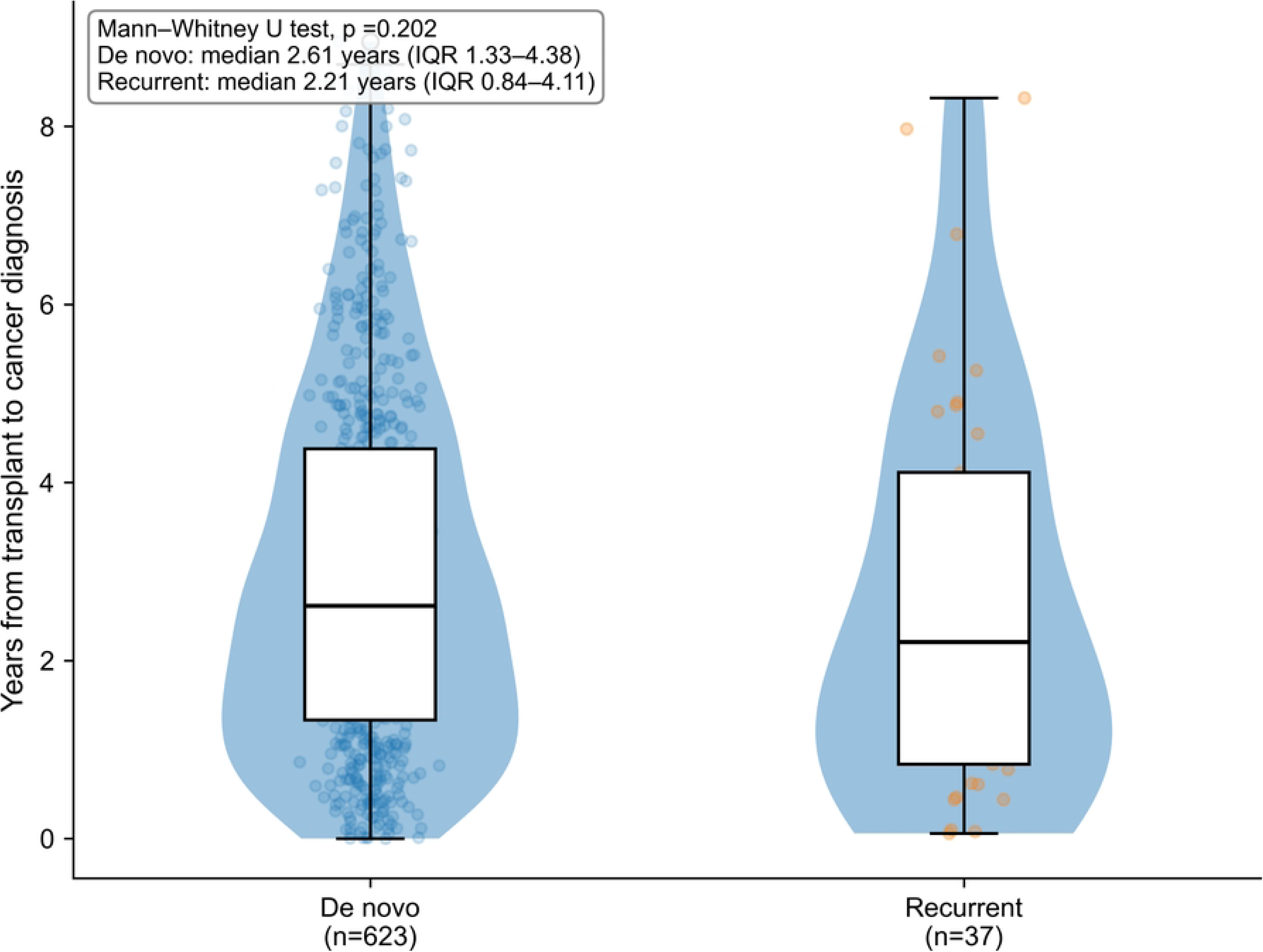
Time from kidney transplantation to prostate cancer diagnosis by phenotype. Violin and box plots show the distribution of time from transplantation to cancer diagnosis among recipients with de novo and recurrent prostate cancer. Boxes represent the interquartile range and horizontal lines indicate medians.

**S2 Fig.**
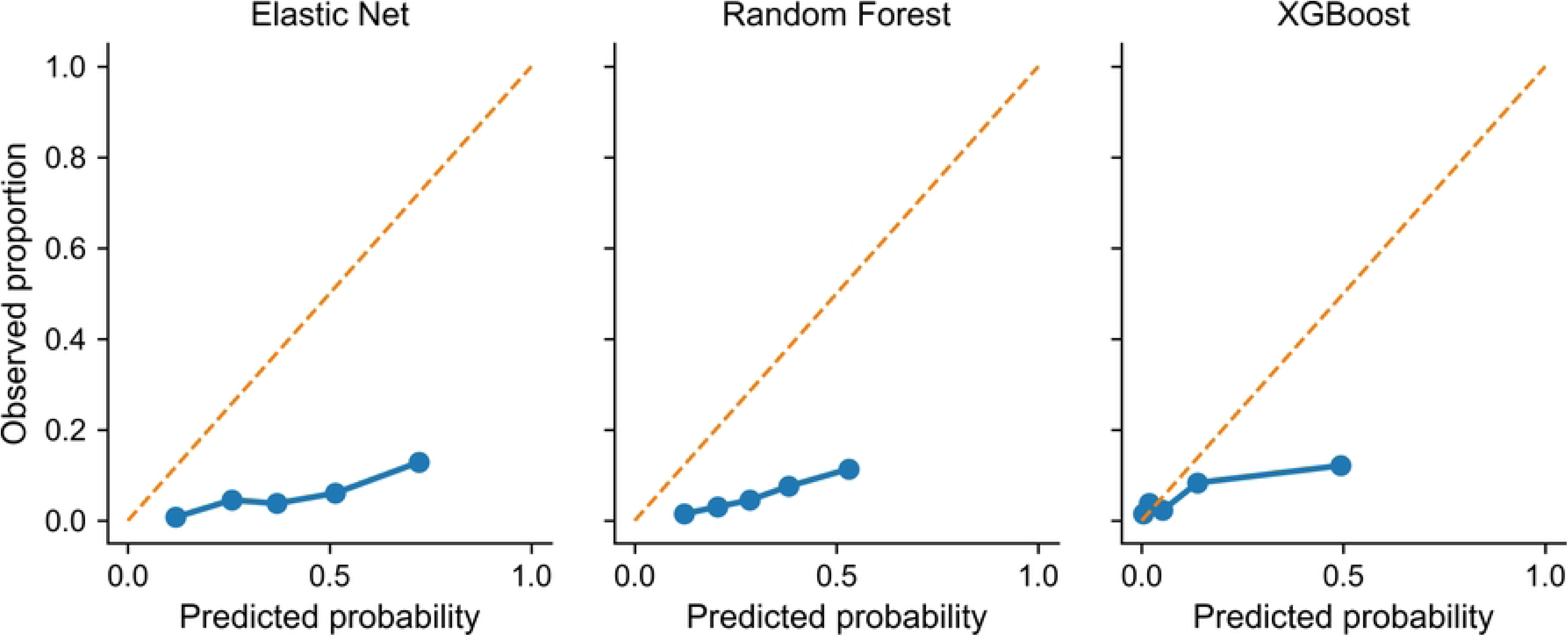
Calibration plots for machine learning models. Calibration plots for elastic net logistic regression, random forest, and XGBoost models using out-of-fold predictions. All models demonstrate overestimation of absolute recurrence risk.

**S3 Fig.**
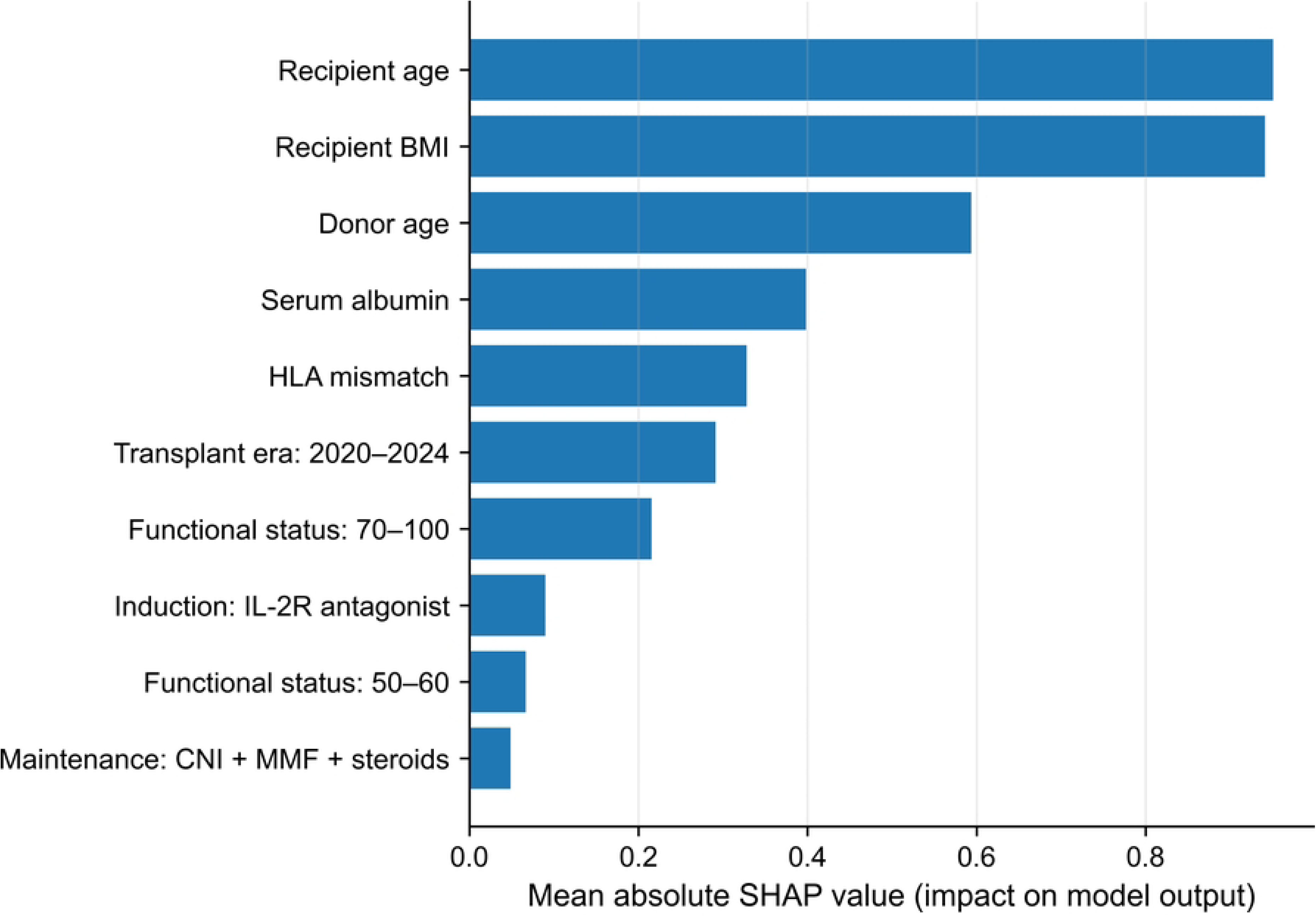
SHAP feature importance for the XGBoost model. Mean absolute SHAP values are shown for the top 10 features, with higher values indicating greater contribution to predicted probability of recurrent prostate cancer phenotype. Functional status categories reflect the collapsed coding used for modeling. SHAP, SHapley Additive exPlanations; BMI, body mass index; CNI, calcineurin inhibitor; HLA, human leukocyte antigen; IL-2R, interleukin-2 receptor; MMF, mycophenolate mofetil.

**S4.** Checklist. STROBE checklist for cohort studies.

## Notes

### Competing Interest Statement

The authors have declared no competing interest.

### Author Declarations

The study received an exemption from the George Mason University Institutional Review Board because it involved secondary analysis of de-identified registry data without access to information that could identify individual participants.

## References

1. Wolfe RA, Ashby VB, Milford EL, Ojo AO, Ettenger RE, Agodoa LYC, et al. Comparison of Mortality in All Patients on Dialysis, Patients on Dialysis Awaiting Transplantation, and Recipients of a First Cadaveric Transplant. N Engl J Med. 1999 Dec 2;341(23):1725–30. doi:10.1056/NEJM199912023412303

2. Ietto G, Gritti M, Pettinato G, Carcano G, Gasperina DD. Tumors after kidney transplantation: a population study. World J Surg Oncol. 2023 Jan 23;21(1):18. doi:10.1186/s12957-023-02892-3

3. Kapoor A. Malignancy in kidney transplant recipients. Drugs. 2008;68 Suppl 1:11–9. doi:10.2165/00003495-200868001-00003 PubMed PMID: 18442297.

4. Massicotte-Azarniouch D, Noel JA, Knoll GA. Epidemiology of Cancer in Kidney Transplant Recipients. Semin Nephrol. 2024 Jan 1; Clinical Innovations in Transplant Onconephrology44(1):151494. doi:10.1016/j.semnephrol.2024.151494

5. Au E, Wong G, Chapman JR. Cancer in kidney transplant recipients. Nat Rev Nephrol. 2018 Aug;14(8):508–20. doi:10.1038/s41581-018-0022-6.

6. Engels EA, Pfeiffer RM, Fraumeni JF, Kasiske BL, Israni AK, Snyder JJ, et al. Spectrum of Cancer Risk Among US Solid Organ Transplant Recipients. JAMA. 2011 Nov 2;306(17):1891. doi:10.1001/jama.2011.1592.

7. Kasiske BL, Snyder JJ, Gilbertson DT, Wang C. Cancer after Kidney Transplantation in the United States. Am J Transplant. 2004 Jun;4(6):905–13. doi:10.1111/j.1600-6143.2004.00450.x.

8. Vajdic CM, McDonald SP, McCredie MRE, Van Leeuwen MT, Stewart JH, Law M, et al. Cancer Incidence Before and After Kidney Transplantation. JAMA. 2006 Dec 20;296(23):2823. doi:10.1001/jama.296.23.2823.

9. Dantal J, Pohanka E. Malignancies in renal transplantation: an unmet medical need. Nephrol Dial Transplant. 2007 May;22 Suppl 1:i4–10. doi:10.1093/ndt/gfm085.

10. Manickavasagar R, Thuraisingham R. Post renal-transplant malignancy surveillance. Clin Med. 2020 Mar;20(2):142–5. doi:10.7861/clinmed.2019-0423.

11. Bray F, Laversanne M, Sung H, Ferlay J, Siegel RL, Soerjomataram I, et al. Global cancer statistics 2022: GLOBOCAN estimates of incidence and mortality worldwide for 36 cancers in 185 countries. CA Cancer J Clin. 2024;74(3):229–63.doi:10.3322/caac.21834.

12. Conti DV, Darst BF, Moss LC, Saunders EJ, Sheng X, Chou A, et al. Trans-ancestry genome-wide association meta-analysis of prostate cancer identifies new susceptibility loci and informs genetic risk prediction. Nat Genet. 2021 Jan;53(1):65– 75. doi:10.1038/s41588-020-00748-0.

13. Damaschke NA, Yang B, Bhusari S, Svaren JP, Jarrard DF. Epigenetic susceptibility factors for prostate cancer with aging. Prostate. 2013 Dec;73(16):1721–30. doi:10.1002/pros.22716.

14. Kratzer TB, Mazzitelli N, Star J, Dahut WL, Jemal A, Siegel RL. Prostate cancer statistics, 2025. CA Cancer J Clin. 2025;75(6):485–97. doi:10.3322/caac.70028.

15. McHugh J, Saunders EJ, Dadaev T, McGrowder E, Bancroft E, Kote-Jarai Z, et al. Prostate cancer risk in men of differing genetic ancestry and approaches to disease screening and management in these groups. Br J Cancer. 2022 Jun;126(10):1366–73. doi:10.1038/s41416-021-01669-3.

16. Rawla P. Epidemiology of Prostate Cancer. World J Oncol. 2019 Apr;10(2):63–89. doi:10.14740/wjon1191.

17. Blosser CD, Haber G, Engels EA. Changes in cancer incidence and outcomes among kidney transplant recipients in the United States over a thirty-year period. Kidney Int. 2021 Jun;99(6):1430–8. doi:10.1016/j.kint.2020.10.018.

18. Wang Y, Lan GB, Peng FH, Xie XB. Cancer risks in recipients of renal transplants: a meta-analysis of cohort studies. Oncotarget. 2018 Mar 16;9(20):15375–85. doi:10.18632/oncotarget.23841.

19. Bao JM, Zhu HL, Yang GS, Chen PL, Dang Q, Chen XX, et al. No significant association between immunosuppression in solid organ transplantation and prostate cancer risk: a meta-analysis of cohort studies. Transl Cancer Res. 2019 Jun;8(3):939– 49. doi:10.21037/tcr.2019.06.03.

20. Bratt O, Drevin L, Prütz KG, Carlsson S, Wennberg L, Stattin P. Prostate cancer in kidney transplant recipients: a nationwide register study. BJU Int. 2020 May;125(5):679–85. doi:10.1111/bju.15002.

21. Kim H, Chae KH, Choi A, Kim MH, Hong JH, Choi BS, et al. Increased risk of genitourinary cancer in kidney transplant recipients: a large-scale national cohort study and its clinical implications. Int Urol Nephrol. 2025 Mar;57(3):715–22. doi:10.1007/s11255-024-04244-w.

22. Haroon UH, Davis NF, Mohan P, Little DM, Smyth G, Forde JC, et al. Incidence, Management, and Clinical Outcomes of Prostate Cancer in Kidney Transplant Recipients. Exp Clin Transplant. 2019 Jun;17(3):298–303. doi:10.6002/ect.2018.0048.

23. Kleinclauss F, Gigante M, Neuzillet Y, Mouzin M, Terrier N, Salomon L, et al. Prostate cancer in renal transplant recipients. Nephrol Dial Transplant. 2008 Jul;23(7):2374–80. doi:10.1093/ndt/gfn008.

24. Sherer BA, Warrior K, Godlewski K, Hertl M, Olaitan O, Nehra A, et al. Prostate cancer in renal transplant recipients. Int Braz J Urol. 2017;43(6):1021–32. doi:10.1590/S1677-5538.IBJU.2016.0510.

25. Spatafora P, Sessa F, Caroassai Grisanti S, Bisegna C, Saieva C, Roviello G, et al. Prostate Cancer Characteristics in Renal Transplant Recipients: A 25-Year Experience from a Single Centre. Front Surg. 2021 Jul 29;8. doi:10.3389/fsurg.2021.716861

26. Sridhar A, Yohannan B, Kaur H, Maithel N. De novo prostate cancer in renal transplant recipients: A single-center study. J Clin Oncol. 2023 Feb 20;41(6 Suppl):326. doi:10.1200/JCO.2023.41.6_suppl.326.

27. Chukwu CA, Wu HHL, Pullerits K, Garland S, Middleton R, Chinnadurai R, et al. Incidence, Risk Factors, and Outcomes of De Novo Malignancy following Kidney Transplantation. J Clin Med. 2024 Mar 24;13(7):1872. doi:10.3390/jcm13071872.

28. Wong G, Lim WH. Prior cancer history and suitability for kidney transplantation. Clin Kidney J. 2023 Nov;16(11):1908–16. doi:10.1093/ckj/sfad141.

29. Heinze G, Schemper M. A solution to the problem of separation in logistic regression. Stat Med. 2002 Aug 30;21(16):2409–19. doi:10.1002/sim.1047.

30. Riley RD, Ensor J, Snell KIE, Harrell FE, Martin GP, Reitsma JB, et al. Calculating the sample size required for developing a clinical prediction model. BMJ. 2020 Mar 18;368:m441. doi:10.1136/bmj.m441.

31. van Smeden M, Moons KG, de Groot JA, Collins GS, Altman DG, Eijkemans MJ, et al. Sample size for binary logistic prediction models: Beyond events per variable criteria. Stat Methods Med Res. 2019 Aug;28(8):2455–74. doi:10.1177/0962280218784726.

32. Leitzmann M, Rohrmann S. Risk factors for the onset of prostatic cancer: age, location, and behavioral correlates. Clin Epidemiol. 2012;4:1–11. doi:10.2147/CLEP.S16747.

33. Zhang S, Lee E, Bopardikar S, Goldstein AS. Defining aging-associated factors that increase susceptibility to prostate cancer. Endocr Relat Cancer. 2025 Sep 8;32(9):e250026. doi:10.1530/ERC-25-0026.

34. Kwak HY, Chae BJ, Bae JS, Jung SS, Song BJ. Breast cancer after kidney transplantation: a single institution review. World J Surg Oncol. 2013 Mar 22;11:77. doi:10.1186/1477-7819-11-77.

35. Hanaway MJ, Woodle ES, Mulgaonkar S, Peddi VR, Kaufman DB, First MR, et al. Alemtuzumab Induction in Renal Transplantation. N Engl J Med. 2011 May 19;364(20):1909–19. doi:10.1056/NEJMoa1009546.

36. Kirk AD, Cherikh WS, Ring M, Burke G, Kaufman D, Knechtle SJ, et al. Dissociation of Depletional Induction and Posttransplant Lymphoproliferative Disease in Kidney Recipients Treated With Alemtuzumab. Am J Transplant. 2007 Nov;7(11):2619–25. doi:10.1111/j.1600-6143.2007.01972.x.

37. Arora VK, Schenkein E, Murali R, Subudhi SK, Wongvipat J, Balbas MD, et al. Glucocorticoid Receptor Confers Resistance to Antiandrogens by Bypassing Androgen Receptor Blockade. Cell. 2013 Dec;155(6):1309–22. doi:10.1016/j.cell.2013.11.012.

38. Austin PC, Lee DS, Wang B. The relative data hungriness of unpenalized and penalized logistic regression and ensemble-based machine learning methods: the case of calibration. Diagn Progn Res. 2024 Nov 5;8(1):15. doi:10.1186/s41512-024-00179-z.

39. Christodoulou E, Ma J, Collins GS, Steyerberg EW, Verbakel JY, Van Calster B. A systematic review shows no performance benefit of machine learning over logistic regression for clinical prediction models. J Clin Epidemiol. 2019 Jun; 110:12–22. doi:10.1016/j.jclinepi.2019.02.004.

